# Episodic future thinking and compassion reduce public health guideline noncompliance urges: A randomised controlled trial

**DOI:** 10.1101/2021.09.13.21263407

**Authors:** Simon van Baal, Antonio Verdejo-García, Jakob Hohwy

## Abstract

**Background:** During the COVID-19 pandemic, public health departments have issued guidelines to limit viral transmission. In this environment, people will feel urges to engage in activities that violate these guidelines, but research on guideline adherence has been reliant on surveys asking people to self-report their typical behaviour, which may fail to capture these urges as they unfold. Guideline adherence could be improved through behaviour change interventions, but considering the wide range of behaviours that COVID-19 guidelines prescribe, there are few methods that allow observing changes of aggregate guideline adherence in the ‘wild’.

**Methods:** In order to administer interventions and to obtain contemporaneous data on a wide range of behaviours, we use ecological momentary assessment. In this preregistered parallel randomised trial, 95 participants aged 18-65 from the UK were assigned to three conditions using blinded block randomisation, and engage in episodic future thinking (n = 33), compassion exercises (n = 31), or a sham procedure (n = 31) and report regularly on the intensity of their occurrent urges (min. 1, max. 10) and their ability to control them. We investigate whether state impulsivity and vaccine attitudes predict guideline adherence, while assessing through which mechanism these predictors affect behaviour.

**Findings:** We found that episodic future thinking (*b* = -1.60) and compassion exercises (*b* = -1.40) reduce the intensity of urges. State impulsivity is associated with stronger urges, but we found no evidence that vaccine hesitancy predicts lesser self-control.

**Interpretation:** We conclude that episodic future thinking exercises and compassion training may be used to stimulate compliance of individuals who are a risk for the community, such as those in voluntary self-isolation.

## Introduction

Enhancing adherence to national COVID-19 health guidelines is essential for saving lives and preventing serious illness. However, studying predictors and methods for improving COVID-19 guideline adherence is challenging because this behaviour is not readily observed in laboratory settings, nor easy to reveal with self-report cross sectional surveys. The focus of this study is, using ecological momentary assessment rather than one-shot surveys, to find whether episodic future thinking and compassion exercises could contribute to increasing adherence to public health guidelines for preventing Covid-19 spread. We also investigate whether state impulsivity and vaccine attitudes predict guideline adherence, while assessing through which mechanism these predictors affect behaviour.

A wealth of research has focused on best-practice for public health communication in order to achieve optimal guideline adherence of the public (1). Adhering to guidelines such as staying home during a lockdown can have immediate personal adverse impact on one’s financial situation (2), mental health (3-5), and physical health (6, 7). In contrast, the effects of non-adherence are often less immediate: when an individual contracts COVID-19 it may take time before symptoms and accompanying negative consequences are experienced; and those consequences may often affect others, such as infecting loved ones, or causing outbreaks in the community. Decisions on guideline adherence therefore constitute a dilemma between choosing the long-term greater good, over the short-term individual gain. Here, we therefore test if increasing people’s future-orientedness, and willingness to make decisions for others’ benefit has potential for increasing adherence.

Episodic future thinking (EFT) refers to imagining or simulating experiences that might occur in one’s personal future, and is known to decrease delay discounting (8, 9). The effects of EFT on delay discounting imply that the valuation of immediate rewards is diminished relative to future rewards. This means that EFT likely affects the intensity of urges, but the effects of EFT in various domains suggest that EFT might also impact self-control (10, 11). Accordingly, this study investigates whether engaging in EFT exercises can change the intensity of urges and self-control.

Adherence to guidelines is ultimately also prosocial, and prosocial behaviour can be enhanced by stimulating compassion, the feeling that arises in witnessing another’s suffering and that motivates a subsequent desire to help (12). Compassion training has a valuation element in addition to a behavioural element, which means compassion training could affect both the intensity of urges and self-control (13, 14); which this study will investigate.

Vaccine hesitancy, attitudes on the effectiveness of these vaccines, and predictions about how soon the pandemic will end could factor into people’s behaviour. These attitudes are usually linked to other attitudes and behaviours relevant to pandemic behaviour such as lesser social distancing and mask wearing (15, 16). We therefore also test how such predictors of guideline adherence influence moment-to-moment behaviour.

Impulsivity, the tendency to make rapid responses for short-term gratification and with insufficient regard for negative consequences (17), is negatively correlated to public health guidelines adherence (18, 19). The excessive future discounting characteristic of impulsivity can be influenced by fluctuations in internal states (20). To understand how impulsivity affects moment-to-moment behaviour, it is important to attain information on people’s mental state when behaviours occur (21, 22). Thus, we study these behaviours in an ecologically valid manner, where proximal information on state impulsivity is obtained.

This study seeks to avoid the distortions that often afflict self-report measures about typical behaviour (23). In order to gain insights into moment-to-moment preventative behaviour in the ‘wild’ and real-time changes following behaviour change interventions, we employ an ecological momentary assessment (EMA; or experience sampling) paradigm. Participants are asked on multiple occasions each day about their urges pertaining to non-adherence of COVID-19 guidelines, and whether they were able to control them.

In this experiment, the main variables of interest are the perceived intensity of urges, and the probability of controlling urges, operationalised as self-control. In our preregistered analyses we predicted that both the compassion intervention and the episodic future thinking intervention would increase the likelihood of controlling urges, where we analyse their effect on self-control and on the intensity of urges. Furthermore, we predicted that state impulsivity is associated with stronger urges and fewer attempts to resist urges, and that vaccine hesitancy and shorter predicted back-to-normal time frames are negatively correlated with the likelihood of controlling an urge through diminishing self-control. In brief, we find that Compassion and EFT interventions reduce the intensity of urges, but we find no evidence that self-control is affected. State impulsivity is significantly related to the intensity of urges.

## Methods

### Participants

The final sample contained 95 participants residing in the UK obtained through volunteer sampling using the Prolific participant recruitment platform. Participants were required to be between 18-65 years of age, and within that age group we created a representative sample, based on sex and age (2×4). Further details on the sample in Appendix A.

In the final sample, there were 40 (42.1%) males, with a mean age of 41.4 (*SD* = 14.1), and 55 females (57.9%) with a mean age of 41.0 (*SD* = 12.5).

40 (42.1%) participants reported they had received a COVID-19 vaccine, and 5 reported that they had been diagnosed with COVID-19 at some point, with one participant reporting they had experienced both of these events. Even though the individual risk of COVID-19 is mitigated for vaccinated individuals and those who previously contracted COVID-19, they were still required to comply with public health guidelines. Therefore, vaccinated individuals were not excluded.

To determine sample size, we estimated an effect size of a 5-percentage point increase in the probability to control an urge in the EFT and Compassion treatments, and assumed that participants would indicate they had an urge 3 times a day. We identified that 95% power could be achieved by collecting data from 90 participants. Code is available <u>here</u>.

### Randomisation and Masking

Participants were randomly assigned a condition according to block randomisation (8 strata, 3 treatments, sequence obtained from sealedenvelope.com), see Figure A1 for the distribution of age and sex per condition. Participants were unaware of their condition assignment, and were either assigned to the EFT condition, the compassion condition, or the control condition.

### Procedure

Participants were asked whether they, or one of their family members, were part of a group that is vulnerable to COVID-19. Participants also answered questions on their willingness to take a COVID-19 vaccine and their beliefs about vaccine efficacy. We also elicited predictions of when people would be able to resume on-site work (insofar as that will ultimately be the case), when people would be able to go on holiday, and when life would go back to ‘normal’. These predictions and vaccination attitudes were then combined into three scores, varying from 0 to 10.

Each morning at 7.30am (expiry time 10am), participants would be asked to do either an EFT exercise, a compassion (i.e., mentalising + affect-matching) exercise, or reflect on a recent news story related to COVID-19 (respectively). All prompts are included in Table A1. After each condition-dependent prompt, participants would be prompted with “Remember that your behaviour has an effect on the COVID-19 situation”. Videos of the user interface can be accessed <u>here</u>.

Throughout each day, participants would receive 5 surveys that were available for 1 hour. In randomised order, they were asked whether since the last survey they had felt an urge to *not* wash their hands, *not* cover their mouths when coughing or sneezing, *not* socially distance (e.g. to hug, shake hands), *not* leave details for contact tracing, or whether they had felt an urge to leave their house, touch their face, or avoid getting tested when it would have been better to do the opposite - from a COVID-19 standpoint. Participants responded using a slider [0,10], where 0 indicated no urge, 1 indicated a very weak urge, and 10 indicated a very strong urge. We then administered the Momentary Impulsivity Scale (24).

The experiment took place from 29 March to 4 April 2021. The UK was in a state of lockdown at that point in time, but most regions in the UK were in the early phase of reopening. Notably, the UK government announced on 29 March that 6 people from 6 different households would be allowed to meet outside. This, together with Easter weekend, produced a situation wherein people were likely to have non-adherence urges. See Figure 1 for a visualisation of urge intensities per day during the experiment.

**Figure 1.**
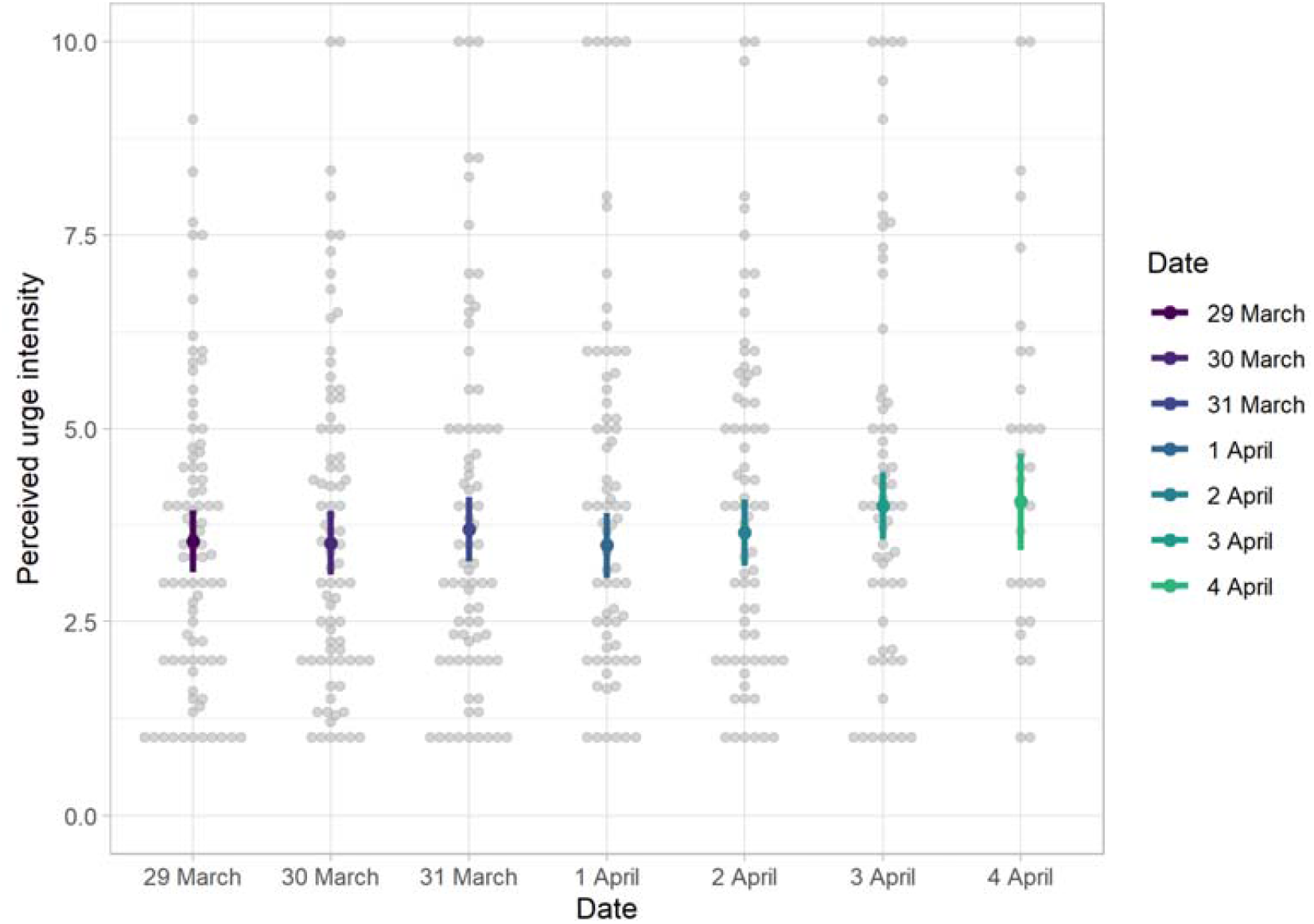
The perceived urge intensity (y-axis), partitioned by day (x-axis). The experiment was partially conducted over a public holiday, at which time stronger urges of non-adherence would be expected. 2 April (third line from the right) was Good Friday, and 4 April (the right-most line) was Easter Sunday. The coloured points represent the estimated marginal means, and the error bars are 95% CIs. The grey data points each represent the aggregated data of one participant.

### Analysis

Our analyses were preregistered, and can be accessed <u>here</u>, data and code can be found <u>here</u>.

The effects of impulsivity on the strength of an urge was analysed using a Linear Mixed Model (LMM), with predictors: condition, the type of urge (and interaction between those), state impulsivity, with the time of day and the day as control variables, and the participant as the random intercept. The type of urge was not included as a predictor in the main preregistered model, but it was specified in the exploratory analyses section and because the effects of the intervention might differ across domains, we decided to include it in the main analyses. The analysis, conducted with the afex package (25) used F-tests of fixed effects terms with Kenward-Roger approximation of degrees of freedom to calculate p-values.

We used a binomial generalised mixed model (GLMM) to conduct the self-control analysis. In addition to the variables in the model above, vaccine hesitancy, vaccine effectiveness beliefs, back-to-normal timeline predictions, and whether participants attempted to resist the urge were included. In order to calculate p-values, likelihood ratio tests were used.

In both analyses, we decided to deviate from the preregistration by including interaction terms with the intervention because we deemed it likely that the intervention might affect some urges more than others. The standard p<.05 criterion was used for determining whether the effects based on the Linear Mixed Model (LMM), or on the Generalised Linear Mixed Model (GLMM) were significantly different from those expected if the null hypothesis were correct. Reported effect sizes are model coefficients for numerical variables, and post-hoc tests were conducted for factors with the emmeans package, which calculates estimated marginal means of the outcome variable for factor levels. We corrected for multiple comparisons using false discovery rate adjusted p-values in our post-hoc tests.

## Results

Different types of urges occurred at different rates, which matters for interpreting the results. Over the one-week-long experiment, 83 out of the 95 participants reported the urge to leave the house at least once, and did so 6.80 times on average (*SD* = 6.09), 79 reported the urge to touch their face (*M* = 7.91, *SD* = 8.14), while 60 reported the urge to not wash their hands (*M* = 5.03, *SD* = 6.11). Only 21 participants reported the urge to not leave their contact details (*M* = 3.86, *SD* = 7.18), and 17 participants reported the urge to avoid getting tested (*M* = 4.82, *SD =* 7.77). 73 Participants reported the urge to disregard social distancing guidelines (*M* = 4.66, *SD* = 4.37), and 33 participants reported the urge to not cover their mouth (*M* = 2.82, *SD* = 4.39).

There was high variance in the number of urges people experienced, and the average number of urges experienced was similar over the different conditions: in the EFT condition, people had 20.0 urges on average (*SD* = 30.2) in the Compassion condition, people had 23.1 urges on average (*SD* = 16.3) and in the Control condition 23.1 (*SD* = 26.7).

### Predictors for Urge Intensity

#### Main Variables of Interest

In accordance with our predictions, the condition factor (EFT, Compassion, Control) had a significant effect on how strong they perceived urges to be, *F*(2, 110) = 7.59, *p* < .001. Post-hoc tests show that participants experienced weaker urges in the EFT condition (estimated marginal mean (*EMM*) = 3.11, 95% CI [2.33, 3.89], *t*(111) = -3.571, *p* < .01), and in the Compassion condition (*EMM* = 3.30, 95% CI [2.51, 4.09], *t*(105) = -3.113, *p* < .01), than in the Control condition (*EMM* = 4.71, 95% CI [3.94, 5.48]). Urge intensity was not significantly different in the EFT condition from the Compassion condition (*t*(113) = .192, *p* = .67). See Figure 2.

**Figure 2.**
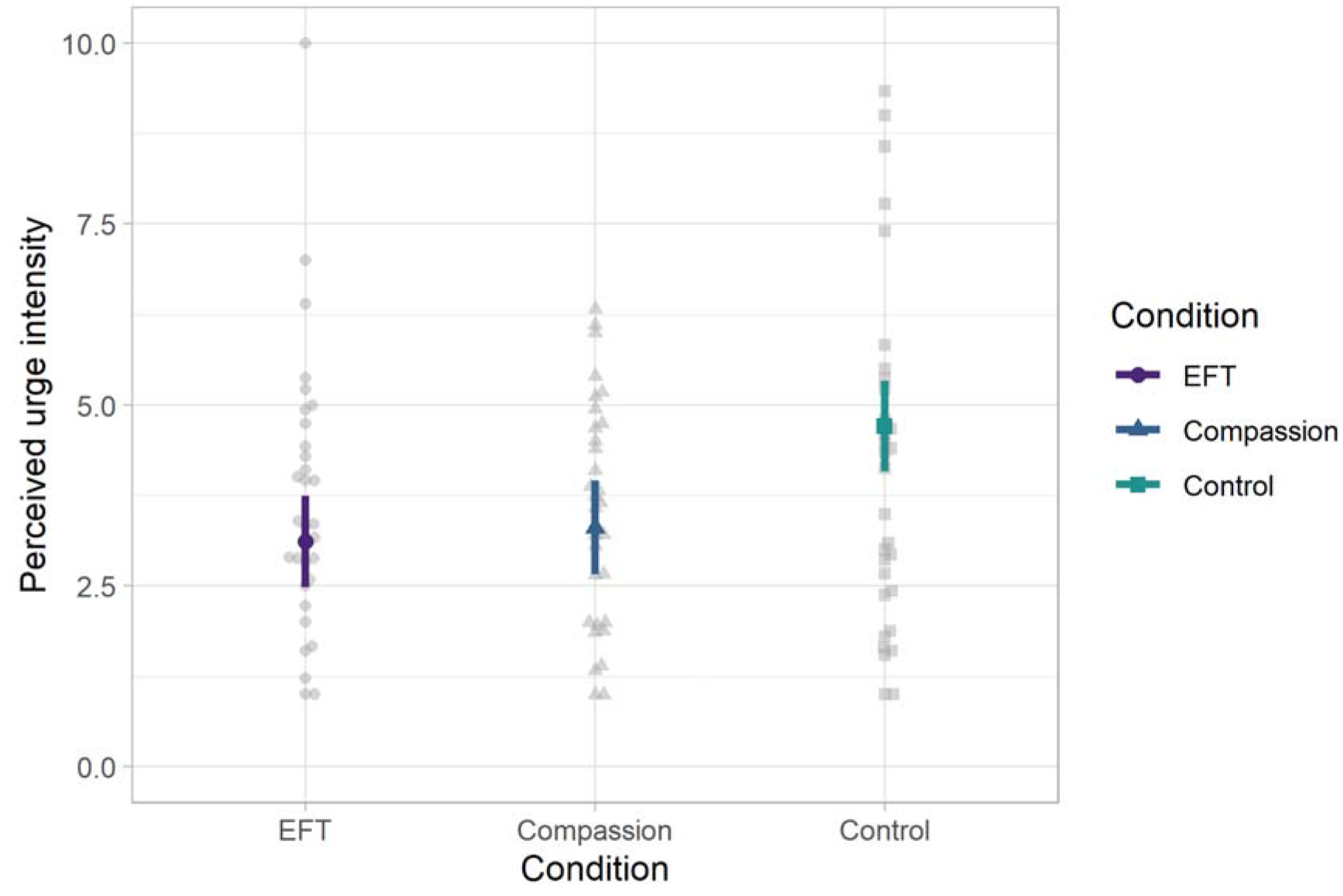
The effects of the between-participants conditions: Episodic Future Thinking (EFT; left, in purple) manipulation, the Compassion manipulation (middle, in blue), as compared to the Control condition (right, in green), on the perceived intensity of urges (y-axis [1,10]). The points are estimated marginal means, and the error bars are 95% CIs. The grey dots each represent the aggregated data of a participant in one of the conditions.

The type of urge had a significant effect on the perceived strength of urges, *F*(6, 2000) = 27.60, *p* < .001. People reported the strongest urges for leaving the house (*EMM* = 4.80, 95% CI [4.42, 5.18]), joint second strongest were urges to avoid getting tested (*EMM* = 3.94, 95% CI [3.34, 4.54]) and urges to not socially distance (*EMM* = 3.94, 95% CI [3.54, 4.35]), fourth were urges to not leave details for contact tracing (*EMM* = 3.78, 95% CI [3.15, 4.40]), fifth were urges to touch one’s face (*EMM* = 3.62, 95% CI [3.24, 4.00]), sixth were urges to avoid washing hands (*EMM* = 3.12, 95% CI [2.70, 3.55]), and the weakest urges pertained to covering mouth and nose when coughing/sneezing (EMM = 2.74, 95% CI [2.19, 3.30]).

The effect of the condition factor on urge intensity also significantly interacted with the type of urge *F*(10, 1999) = 14.36, *p* < .001. Post-hoc tests revealed that urges to avoid leaving details for contact tracing were weaker in the EFT condition (*EMM* = 1.85, 95% CI [.44, 3.26]; *t*(678) = 7.025, *p* < .001), and in the Compassion condition (*EMM* = 2.47, 95% CI [1.00, 3.87]; *t*(644), = 6.161, *p* < .001), than in the Control condition (*EMM* = 7.04, 95% CI [5.96, 8.12]), but were not significantly different from each other, *t*(926) = .695, *p* = .49. Further, urges to avoid getting tested were also weaker in the EFT condition (*EMM* = 1.77, 95% CI [.48, 3.06]; *t*(635) = 7.768, *p* < .001), and in the Compassion condition (*EMM* = 2.68, 95% CI [1.33, 4.02]; *t*(618) = 7.768, *p* < .001), than in the Control condition (*EMM* = 7.38, 95% CI [6.21, 8.55]), but were not different from each other, *t*(744) = 1.170, *p* = .24.

State impulsivity had a significant effect on the perceived strength of urges, *b* = .429, 95% CI [0.262, .597], *F*(1, 2030) = 25.25, *p* < .001. See Figure 3 for a depiction of the relationship between state impulsivity and the intensity of urges.

**Figure 3.**
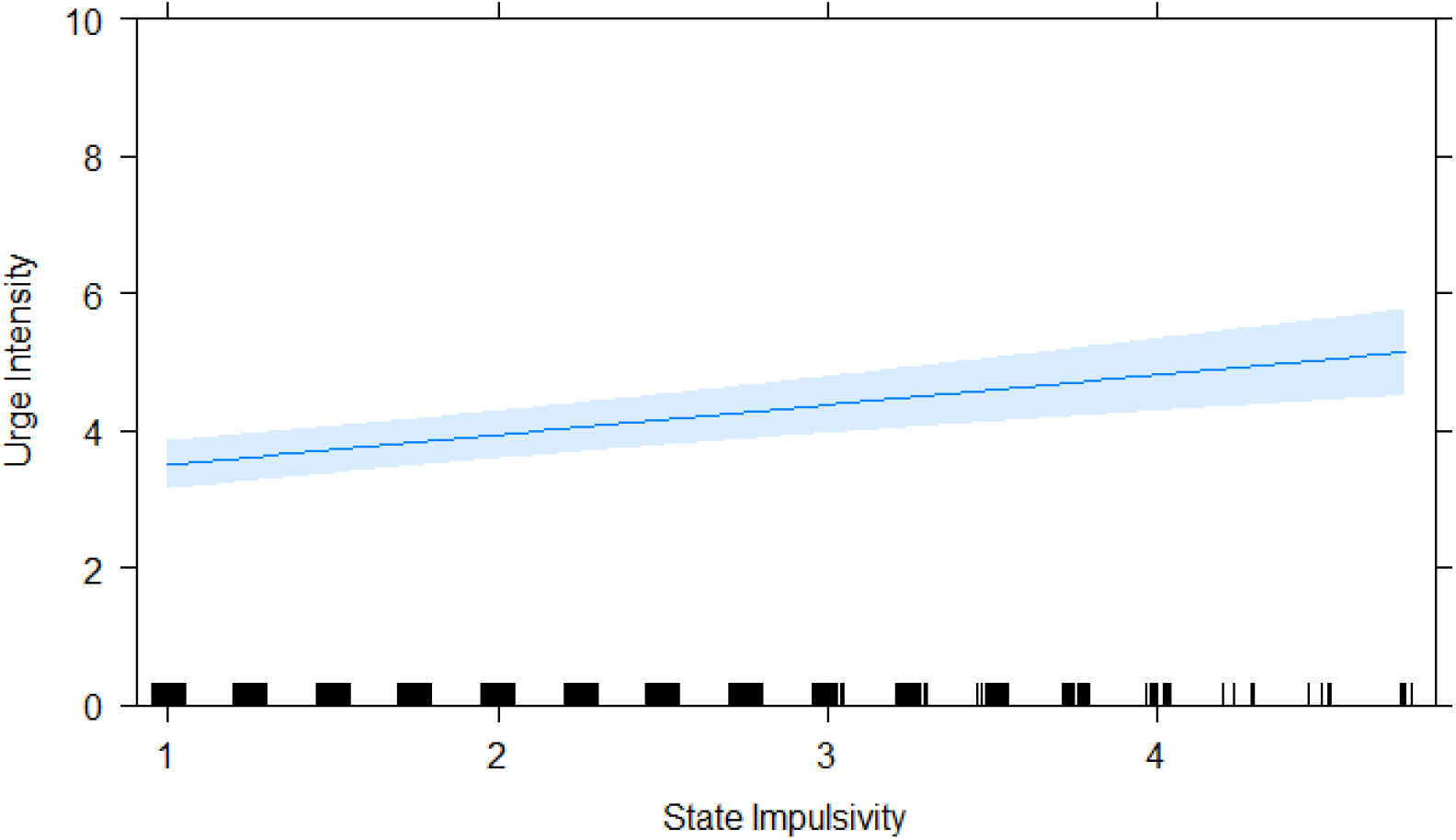
The relationship between state impulsivity as measured by the MIS (x-axis) and the intensity of urges [1,10] (y-axis). The black lines at the bottom represent participants’ MIS scores per filled out survey, and the blue band shows the 95% CI.

### Predictors for Self-Control

The condition factor did not have a significant effect on self-control, χ^2^(2) = 4.56, *p* = .10.

The type of urge had a significant effect on self-control, χ^2^(6) = 219.89, *p* < .001. The estimated marginal means show that people were most likely to control an urge to *not* cover their mouths (*EMM* (Probability) = .690, 95% CI [.498, .833]; they were second most likely to control to *not* wash their hands (*EMM* = .637, 95% CI [.519, .741]); third were urges to *not* socially distance (*EMM* = .517, 95% CI [.407, .625]); fourth were urges to leave the house (*EMM* = .405, 95% CI [.318, .498]); fifth were urges to touch one’s face (*EMM* = .154, 95% CI [.109, .213]), sixth were urges to *not* leave contact details for contact tracing purposes (*EMM* = .086, 95% CI [.036, .193]); the most difficult urges to control were those that pertain to *not* get tested (*EMM* = .009, 95% CI [.003, .031]). It is important to note that these probabilities must be judged together with the relative frequencies of each urge. For instance, the probability of controlling an urge that pertains to avoiding getting tested is extremely low, but only 21 participants ever got one of these urges, for a total of 82 urges. See Figure 4 for a depiction of the probability to control various urges.

**Figure 4.**
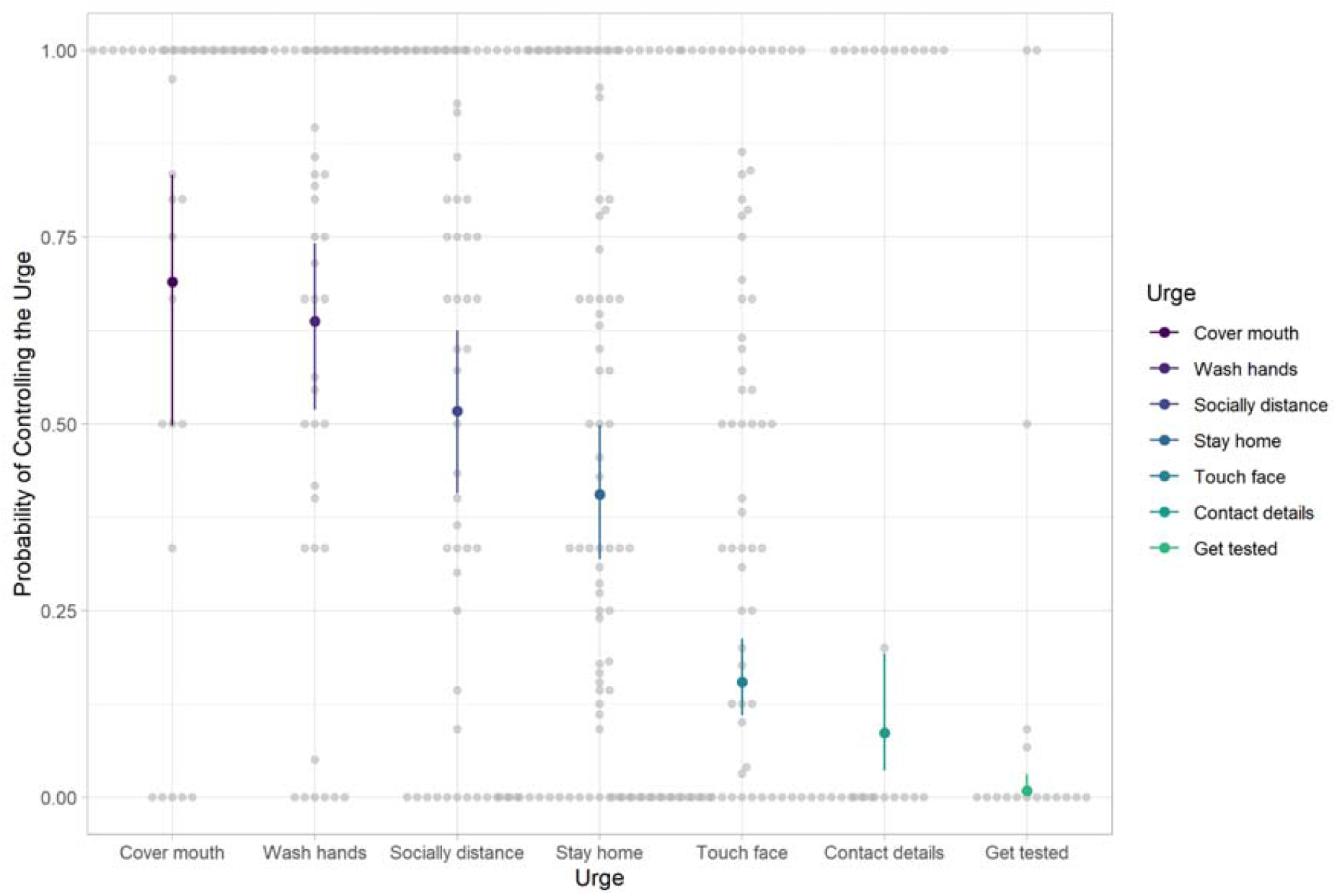
The estimated marginal means for controlling various urges. Each point represents the probability of controlling an urge pertaining to a certain type of behaviour, from left to right: the probability of controlling an urge to not cover one’s mouth and nose when coughing or sneezing, to not wash hands, to not socially distance, to not stay home, to avoid providing contact details when entering an establishment, and to avoid getting tested. The error bars are 95% confidence intervals.

The condition participants were assigned to significantly interacted with the type of urge in predicting self-control, χ^2^(12) = 29.45, *p* < .01.

State impulsivity did not have a significant effect on self-control, *b* = .054, 95% CI [-.224, .332], χ^2^(1) = .16, *p* = .69. Furthermore, state impulsivity did not have a significant effect on the probability of attempting to resist an urge *b* **= -**.108, 95% CI.

Vaccine hesitancy did not have a significant effect on self-control, *b* = -.103, 95% CI [-.270, .063], χ ^2^ (1) = 1.37, *p* < .24. Judgements about vaccine hesitancy did not have a significant effect on self-control (*b* = .038, 95% CI [-.177, .253]; χ^2^(1) = .09, *p* = .77), and neither did predictions about when life would go back to ‘normal’ after the pandemic (*b* = -.141, 95% CI [-.786, .505]; χ^2^(1) = .20, *p* = .66).

## Discussion

Our findings show that episodic future thinking and compassion exercises reduced the intensity of certain urges, but did not affect self-control. This finding broadly aligns with the evidence that EFT can enhance future-oriented decision making in various contexts (10, 11). Given that urges usually pertain to immediate rewards, this reduction in the strength of urges after an EFT exercise is most likely because EFT reduces the relative value of immediate rewards compared to future rewards (8, 9). This finding suggests that an invitation to engage in EFT and compassion-inducing talking points could be incorporated into press conferences and some public announcements to decrease urges of noncompliance during public health crises. People who pose a specific risk to the community (e.g., those in voluntary self-isolation after travelling abroad) could be invited to periodically perform such a task.

The mechanisms through which EFT and compassion exercises affect behaviour differ: EFT is known to enhance future-oriented decision making by decreasing delay discounting (8), whereas compassion exercises can increase altruism (12). These mechanisms both likely promote the salience of the potential negative consequences to people’s actions, which may be the reason for their effectiveness in this context. Alternative explanations include that compassion exercises can lead to increased positive affect and motivation (26), and can be helpful to deal with daily stressors (27), which may also decrease the perceived intensity of urges.

We found no evidence that vaccine attitudes or predictions of back-to-normal timelines were associated with self-control. Other studies show that vaccine hesitancy is correlated with lesser social distancing and mask wearing (16), but most studies reporting these relationships rely on judgements about typical behaviour, or intentions to comply with guidelines. The lack of a significant relationship between vaccine attitudes and guideline adherence in this study suggests that more research is needed to understand how these attitudes affect moment-to-moment behaviour.

We find that different urges occur at varying rates within and between participants, which is an important consideration when assessing the relative impact of these urges. If an urge is relatively rare, but difficult to control, then it may not be as relevant for policymakers and other key stakeholders to see if the motivation to control this urge can be increased. Urges to avoid getting tested, or to not leave contact details were relatively infrequent, but the fact that around 20% of the sample reported one of these urges at least once is worrying given their importance to pandemic management (28).

State impulsivity was related to stronger urges, but not to diminished self-control. This evidence suggests that state fluctuations in impulsivity play an important, but poorly understood role in determining public health guideline adherence during pandemics. A recent study has found this ‘bottom-up’ effect of state impulsivity for a different, more general domain of urges (29). It also suggests that interventions targeting the internal state of the individual, and impulsivity in particular, might be effective at ameliorating their guideline adherence. Future research could investigate whether state impulsivity, as well as other internal states, can be targeted to improve public health guideline adherence.

The main limitations of this study were that the heterogeneity of the experience of certain types of urges rendered the sample size too small to draw accurate inferences in some domains. Only around 20 individuals reported having urges to avoid to getting tested or to avoid leaving details for contact tracing at least once, and these 20 individuals were spread over the 3 conditions. This speaks to the strengths and weaknesses of our ecological momentary assessment paradigm because, on the one hand, it is a powerful paradigm for events that occur often and to a wide range of people (such as the urge to abandon social distancing), and it can provide insight into behaviour ‘in the wild’. On the other hand, for events that only happen for a narrow subset of people, or that happen infrequently, ecological momentary assessment needs to be applied to that particular subset, or another approach should be considered. The other limitation is that it is unclear to what extent the findings apply to different populations.

Future research could focus on the role of behavioural science interventions, including, but not limited to, episodic future thinking and compassion-inducing exercises, in producing desired behaviour during public health crises. We also deem it important that more research is devoted to uncovering the factors predicting moment-to-moment decision making in people’s daily lives, where ecological momentary assessment and GPS data (30) could play a critical role. These methods can provide a more comprehensive understanding of the effects of behavioural interventions, shedding light on their longevity and externalities. For proper understanding of behavioural drivers during the pandemic, there needs to be more emphasis on the proximal predictors to guideline compliance.

## Data Availability

All data and analysis code are available, links are provided in the manuscript. We also provide videos of the data collection procedure.

https://osf.io/pj9ft/

https://osf.io/x5edw/

## Acknowledgements

The authors would like to thank Dr. Peter Koval for the use of the SEMA3 platform.

## Availability of data and materials

### Appendix A

To become eligible for the study, participants (n = 293) completed an eligibility survey prior to the experiment, which was used to create a representative sample. Participants were added to the EMA software in two rounds in order to cope with varying drop-out across the eight groups, since there was more dropout than anticipated during the EMA app download-phase. In total, 200 participants were added to the EMA software, 112 of whom downloaded the app, and 97 of those completed more than 50% of the surveys. As indicated in our preregistration, participants who completed less than 50% of surveys were excluded, the average survey completion rate for the rest was 83.7%. Finally, two participants never reported having urges of non-adherence, and thus these were included from the final sample, given that there was no variation in the data.

Differing dropout across groups after the second round of participant entries led to an imbalance in the female/male sample division. Given that this study is the first of its kind, we cannot assume that these responses are missing at random. In order to address this issue, we analysed whether age and sex influence urges and self-control.

**Figure A1.**
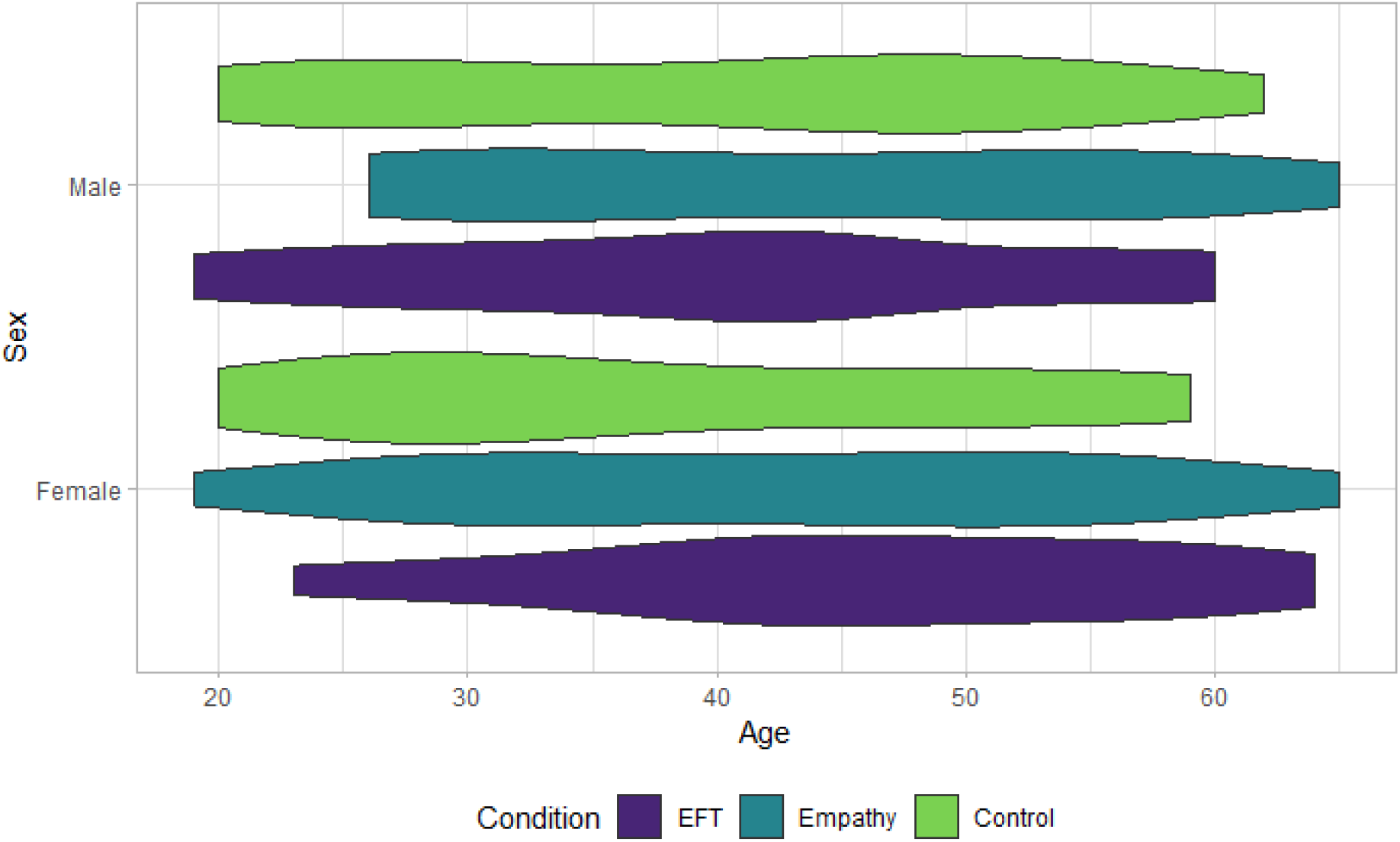
The age and sex distribution of the sample.

### Appendix B

**Table A1.**
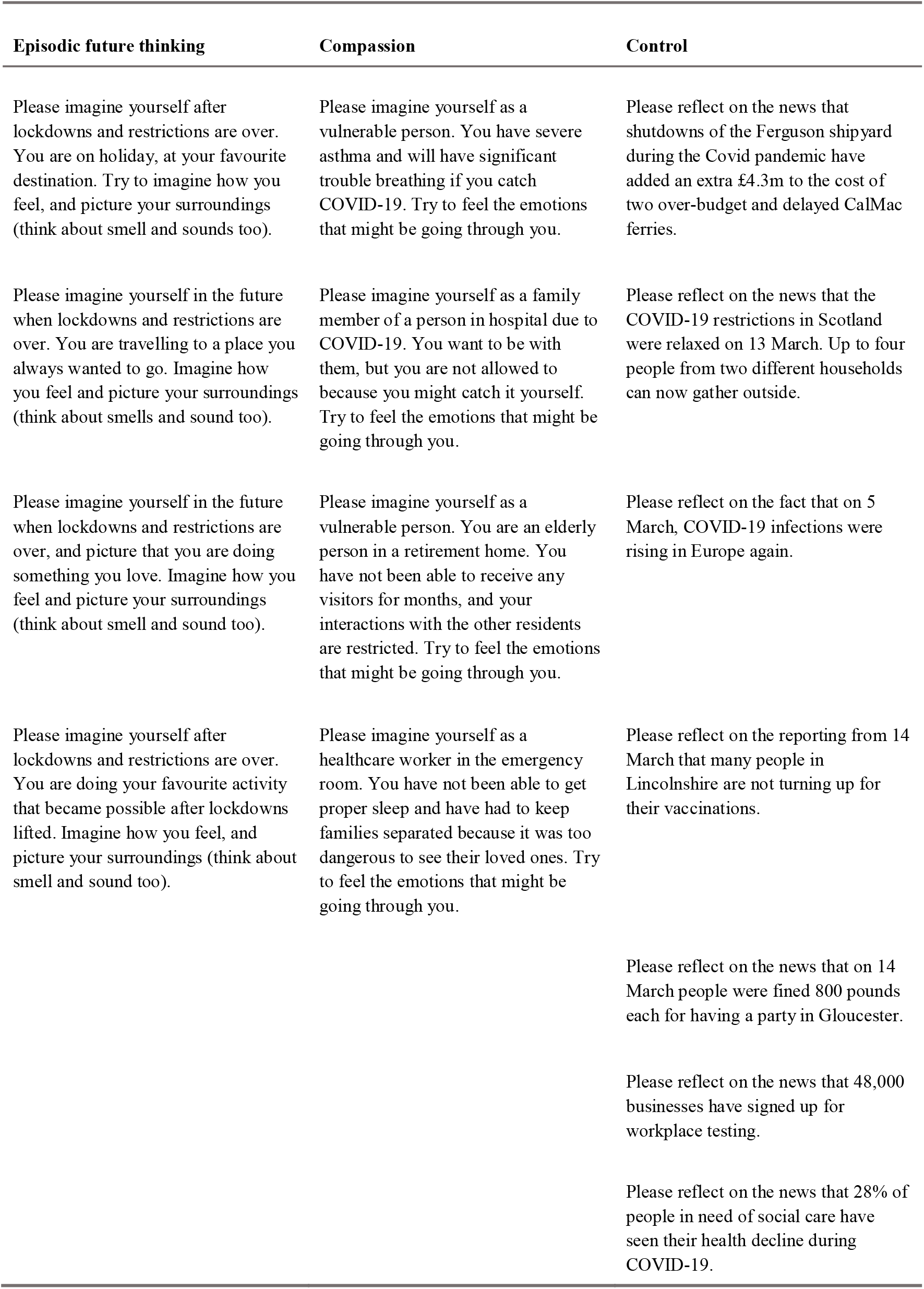
The instructional prompts participants received each morning in their 7.30am survey.

